# Establishing a Bidirectional Correspondence Table between the Japanese Standard Tables of Food Composition 2020 (8th Edition) and the USDA FoodData Central Using Large Language Model-Based Matching

**DOI:** 10.64898/2026.05.10.26352824

**Authors:** Shin-ichi Nakagawa, Akira Yamamoto

## Abstract

**Background:** No official correspondence table exists between the Japanese Standard Tables of Food Composition 2020 (8th edition; MEXT) and the USDA FoodData Central (FDC), despite their widespread use in nutritional research. This absence has hindered international comparison of food composition data for over six decades.

**Methods:** We developed a bidirectional matching pipeline using Claude Haiku (Anthropic), a large language model (LLM), combining food category mapping, 17-nutrient Euclidean distance ranking, and LLM-based conceptual judgment. Survey (FNDDS) data were excluded from FDC, yielding 8,158 items (Foundation Foods and SR Legacy). Matching was performed in both directions: MEXT→FDC and FDC→MEXT.

**Results:** Of 2,478 MEXT items, 1,927 (77.8%) were matched to FDC items, while 549 (22.2%) had no FDC equivalent (JP-only foods). Of 8,158 FDC items, 5,445 (66.7%) were matched to MEXT items, while 2,698 (33.1%) had no MEXT equivalent (US-only foods). Bidirectional consensus yielded 435 confirmed food pairs across 13 food categories. Notably, FDC items showed systematically higher calcium (+6.0 mg/100g) across 12 of 13 categories, while MEXT items showed systematically higher potassium (−3.7 mg/100g) across 9 of 13 categories and higher vitamin A as RAE (−3.7 μg/100g) across 8 of 13 categories.

**Conclusions:** This study presents the first systematic bidirectional food correspondence table between MEXT and USDA FDC. The 435 confirmed pairs constitute a validated common vocabulary for international food composition research. The systematic cross-national differences in calcium, potassium, and vitamin A represent novel findings with direct implications for international dietary comparison studies. The complete correspondence table (Version 0.1) is openly available at https://github.com/shnkgw-rincom/jbfd-correspondence-table (DOI: 10.5281/zenodo.20103327).

## 1. Introduction

Food composition databases are essential tools in nutritional epidemiology, dietary assessment, and public health policy. The two most widely referenced national databases — the Standard Tables of Food Composition in Japan 2020 (8th edition; MEXT) [1] and the USDA FoodData Central (FDC) [2] — each cover thousands of foods with detailed nutrient profiles. However, no official correspondence table between these two databases has ever been established, a gap that has persisted for over six decades since the Japanese tables were first published in 1950.

This absence has significant practical consequences. Researchers conducting cross-national dietary comparisons must rely on ad hoc manual matching, which is labor-intensive, non-reproducible, and prone to conceptual errors. International initiatives such as FAO/INFOODS have established nutrient naming standards (tagnames) [3], but have not produced a comprehensive item-level correspondence between national food composition tables. The Japanese tables also lack an official English translation, further hindering international use.

Recent advances in large language models (LLMs) offer a new approach to this problem. LLMs can reason about food concepts across languages and cultural contexts, enabling semantic matching beyond simple string comparison or nutrient distance alone. Previous work by our group demonstrated that LLM-based matching using nutrient profile vectors significantly improved cross-national food composition database compatibility [4, 5].

In the present study, we applied a bidirectional LLM-based matching pipeline to establish food-item correspondences between MEXT and USDA FDC. Beyond constructing the correspondence table itself, we report a novel finding: systematic, category-consistent differences in calcium, potassium, and vitamin A between matched Japanese and American food items, which we interpret as reflecting genuine structural differences in the food supply of the two countries. Japan-specific foods, US-specific foods, and the interpretation of one-directional matches will be addressed in subsequent papers of this series.

## 2. Materials and Methods

### 2.1 Data sources

The Japanese Standard Tables of Food Composition 2020 (8th edition; MEXT) were obtained from the Ministry of Education, Culture, Sports, Science and Technology, Japan [1]. The dataset comprises 2,478 food items across 18 major food categories, with nutrient values expressed per 100 g edible portion.

The USDA FoodData Central (FDC) database was accessed via the official download portal [2]. For this study, only Foundation Foods (365 items) and SR Legacy (7,793 items) were used (total: 8,158 items). Survey (FNDDS; 5,432 items) was excluded because it comprises composite foods and recipes reflecting dietary survey conditions rather than individual food items with independent composition profiles (Figure 1).

**Figure 1.**
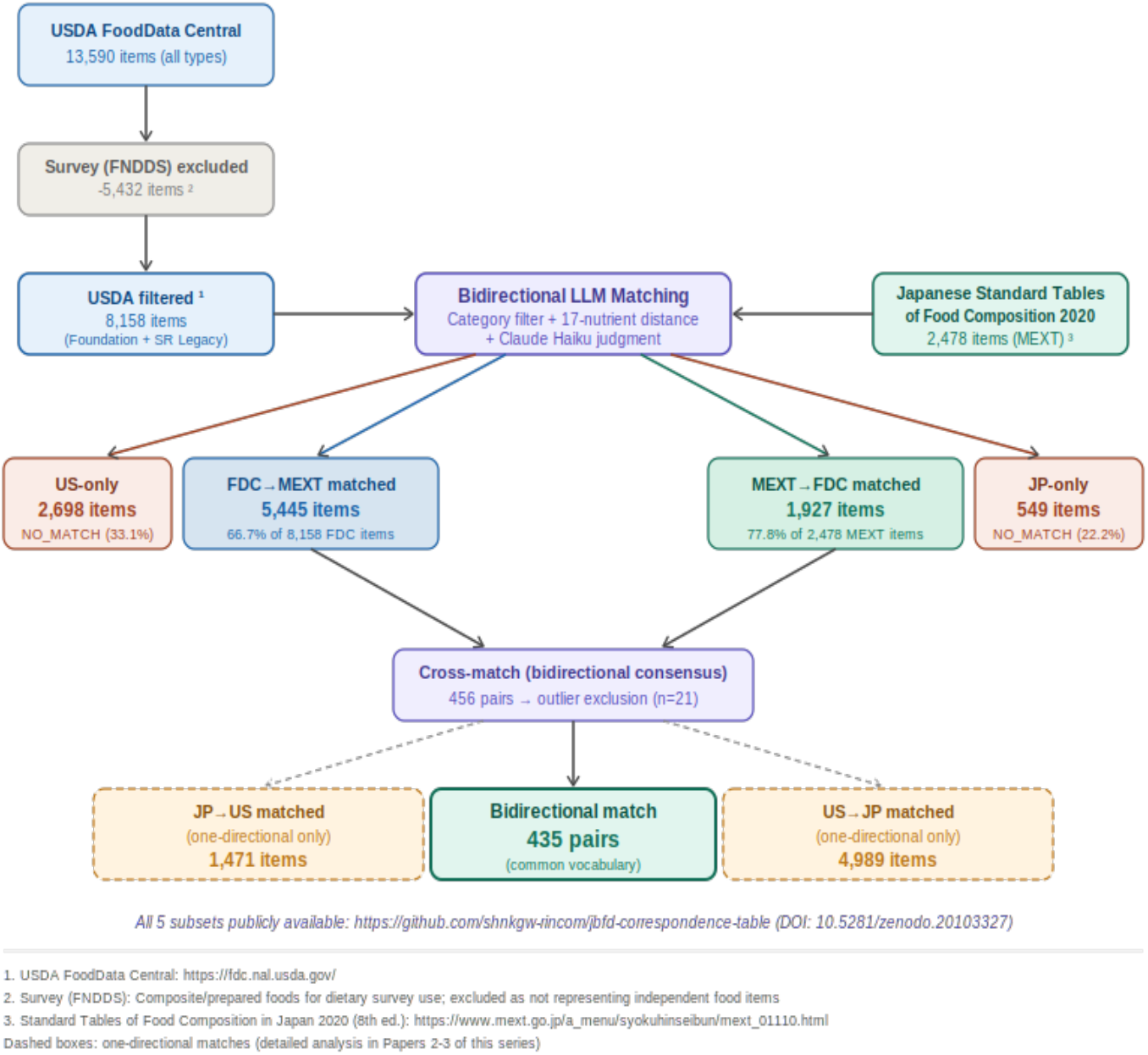
Data processing workflow for bidirectional LLM-based food matching between MEXT and USDA FoodData Central. USDA Survey (FNDDS) data were excluded prior to matching. ^1^https://fdc.nal.usda.gov/ ^2^Composite/prepared foods for dietary survey use ^3^ https://www.mext.go.jp/a_menu/syokuhinseibun/mext_01110.html

### 2.2 Nutrient variables

Seventeen nutrient variables common to both databases were used: energy (kcal), protein (g), fat (g), carbohydrate (g), dietary fiber (g), calcium (mg), iron (mg), potassium (mg), zinc (mg), vitamin C (mg), vitamin A as RAE (μg), vitamin B1 (mg), vitamin B2 (mg), vitamin D (μg), vitamin B12 (μg), and salt equivalent (g). All values are expressed per 100 g edible portion.

### 2.3 Matching pipeline

The matching pipeline consisted of three sequential steps. Step 1 — Category pre-filtering: a manual mapping table was constructed between MEXT major food categories and corresponding FDC food categories. Step 2 — Nutrient distance ranking: for each source item, candidate items in the target category were ranked by Euclidean distance over the 17 normalized nutrient variables, and the top 25 candidates were selected.

Step 3 — LLM judgment: Claude Haiku (claude-haiku-4-5-20251001; Anthropic) was queried with a structured prompt presenting the source food item and 25 candidates, instructed to select the single best conceptual and nutritional match, or to designate NO_MATCH if no adequate equivalent existed. Responses were constrained to JSON format. Matching scripts are publicly available at https://github.com/shnkgw-rincom/jbfd-correspondence-table.

Bidirectional matching was performed independently in both directions: MEXT→FDC and FDC→MEXT. A food pair was designated a bidirectional consensus match if the MEXT item selected the FDC item and the FDC item reciprocally selected the same MEXT item (Figure 2). This process yielded five distinct subsets: bidirectional consensus pairs, MEXT→FDC matched (one-directional), FDC→MEXT matched (one-directional), JP-only foods (MEXT NO_MATCH), and US-only foods (FDC NO_MATCH); all five subsets are publicly available (see Data Availability).

**Figure 2.**
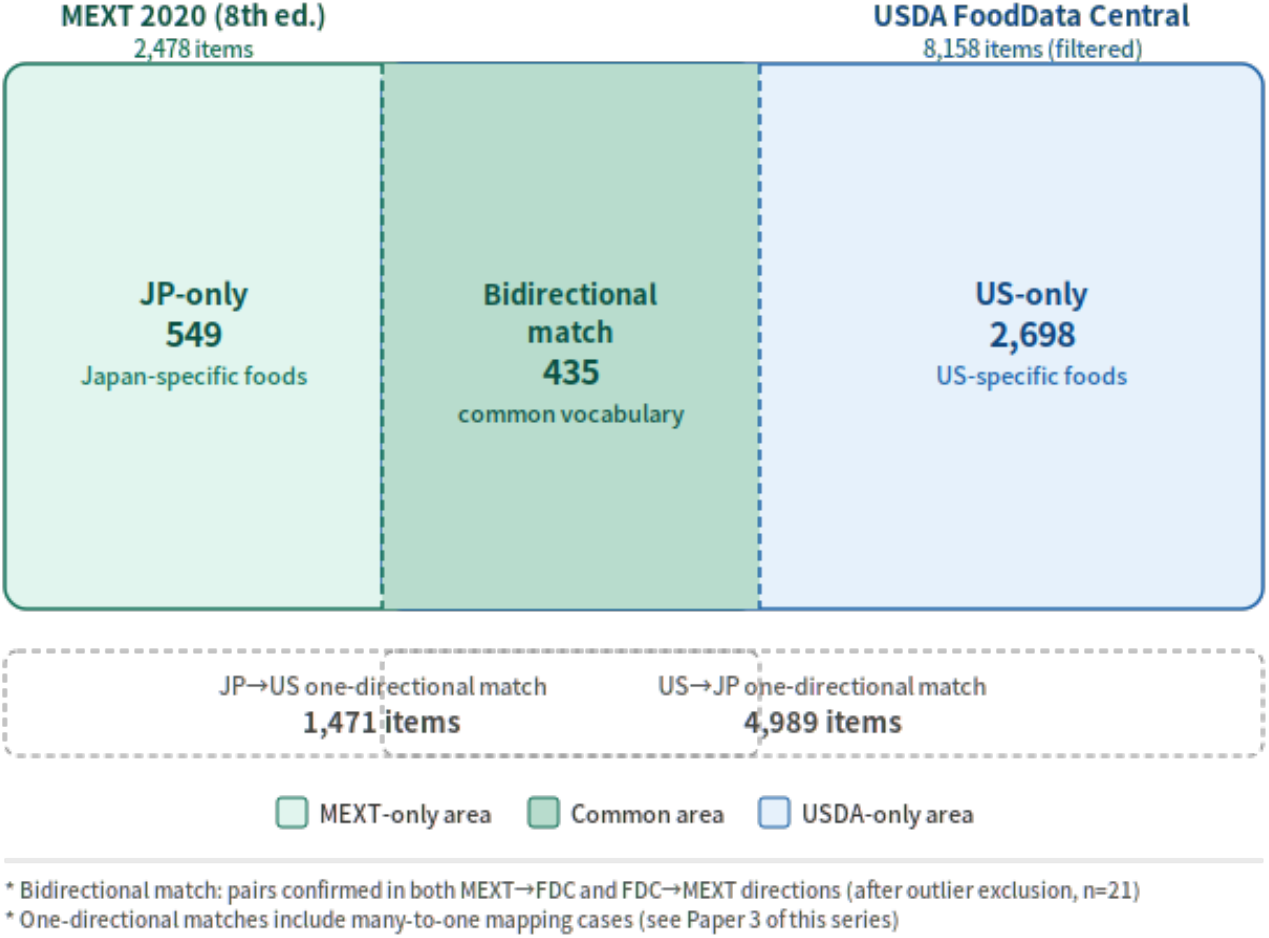
Rectangular Venn diagram illustrating the distribution of food items across MEXT and USDA FoodData Central following bidirectional LLM-based matching. Numbers reflect final counts after outlier exclusion (n=21). Dashed rectangles indicate one-directional match subsets.

### 2.4 Outlier exclusion

Among 456 initial bidirectional consensus pairs, 21 pairs were excluded as outliers based on implausibly large nutrient differences: |ΔEnergy| > 300 kcal, |ΔCa| > 200 mg, |ΔK| > 300 mg, or |ΔVitamin A| > 500 μg per 100 g. The final analysis dataset comprised 435 bidirectional consensus pairs.

### 2.5 Statistical analysis

Nutrient differences (USDA FDC − MEXT) were computed for each of the 17 variables in each matched pair. Category-stratified means were calculated for the 435 confirmed pairs. All analyses were performed using Python 3.11.

## 3. Results

### 3.1 Matching outcomes

Of 2,478 MEXT items, 1,927 (77.8%) were matched to at least one FDC item in the MEXT→FDC direction, while 549 items (22.2%) were designated NO_MATCH (JP-only foods). Of 8,158 FDC items, 5,445 (66.7%) were matched to at least one MEXT item in the FDC→MEXT direction, while 2,698 items (33.1%) were designated NO_MATCH (US-only foods). Bidirectional consensus yielded 456 confirmed pairs, of which 21 were excluded as outliers, leaving 435 pairs for analysis. The overall distribution is shown in Figures 1 and 2.

In addition, 1,471 MEXT items showed one-directional matches (JP→US only) and 4,989 FDC items showed one-directional matches (US→JP only). The large number of US→JP one-directional matches primarily reflects the many-to-one mapping arising from FDC’s greater item granularity; interpretation will be addressed in a subsequent paper.

### 3.2 Category distribution of bidirectional matches

The 435 bidirectional consensus pairs spanned 13 of 18 MEXT major food categories (Table 1). Meat and poultry accounted for the largest share (n = 113, 26.0%), followed by fish and shellfish (n = 53, 12.2%), vegetables (n = 46, 10.6%), and fruits (n = 45, 10.3%). Five MEXT categories — seaweeds, mushrooms, starchy roots/starches, sugars/sweeteners, and eggs — yielded no bidirectional consensus pairs, reflecting the Japan-specific nature of these food groups. Representative matched pairs are shown in Table 2.

**Table 1.** Mean nutrient differences (USDA FoodData Central − MEXT, per 100 g edible portion) for the 435 bidirectional consensus food pairs, stratified by MEXT food category. Shaded columns (ΔCa, ΔK, ΔVit A) highlight key findings. Positive values: FDC higher; negative values: MEXT higher. Outlier pairs (n=21) excluded.

| Category | n | ΔEnergy (kcal) | ΔProtein (g) | ΔFat (g) | ΔCarb (g) | ΔCa (mg) | ΔK (mg) | ΔVit A (μg) |
| --- | --- | --- | --- | --- | --- | --- | --- | --- |
| Meat and poultry | 113 | +0.3 | +0.4 | -0.2 | -0.8 | +6.2 | -13.7 | +5.8 |
| Fish and shellfish | 53 | +0.8 | -0.3 | +0.1 | +1.1 | +2.5 | +20.5 | +41.6 |
| Vegetables | 46 | +11.4 | -0.1 | +0.6 | +2.1 | +6.4 | -19.5 | -51.5 |
| Fruits | 45 | -1.5 | +0.1 | -0.1 | +3.4 | +5.8 | -4.5 | -40.2 |
| Cereals and grains | 43 | +4.7 | -0.4 | -0.0 | +2.6 | +10.2 | +15.7 | -14.3 |
| Dairy products | 30 | +1.6 | -0.3 | -0.4 | +3.6 | +9.4 | +10.5 | +39.3 |
| Beverages | 23 | +8.0 | -0.1 | -1.2 | +0.7 | -2.6 | -3.0 | 0.0 |
| Nuts and seeds | 20 | -3.7 | -0.1 | +1.3 | +9.3 | +5.5 | -55.8 | -35.2 |
| Confectioneries | 18 | -9.6 | -0.5 | -0.8 | -1.7 | -11.8 | +16.9 | +11.5 |
| Legumes | 13 | -5.6 | -1.0 | -0.5 | +4.9 | +17.2 | -17.0 | -21.4 |
| Seasonings/spices | 12 | +29.1 | -0.4 | -0.5 | +5.5 | +10.2 | -13.2 | -8.9 |
| Fats and oils | 11 | -3.7 | -0.1 | +2.0 | -0.5 | -11.0 | +1.0 | 0.0 |
| Processed foods | 8 | +0.3 | +3.1 | -1.0 | -1.2 | +27.8 | +42.9 | 0.0 |
| **Total / Mean** | **435** | **+2.2** | **0.0** | **0.0** | **+2.6** | **+6.0** | **-3.7** | **-3.7** |

Note: $\Delta$ = USDA FDC value – MEXT value. Vit A expressed as retinol activity equivalents (RAE). Salt equivalent = sodium (g) × 2.54.

**Table 2.**
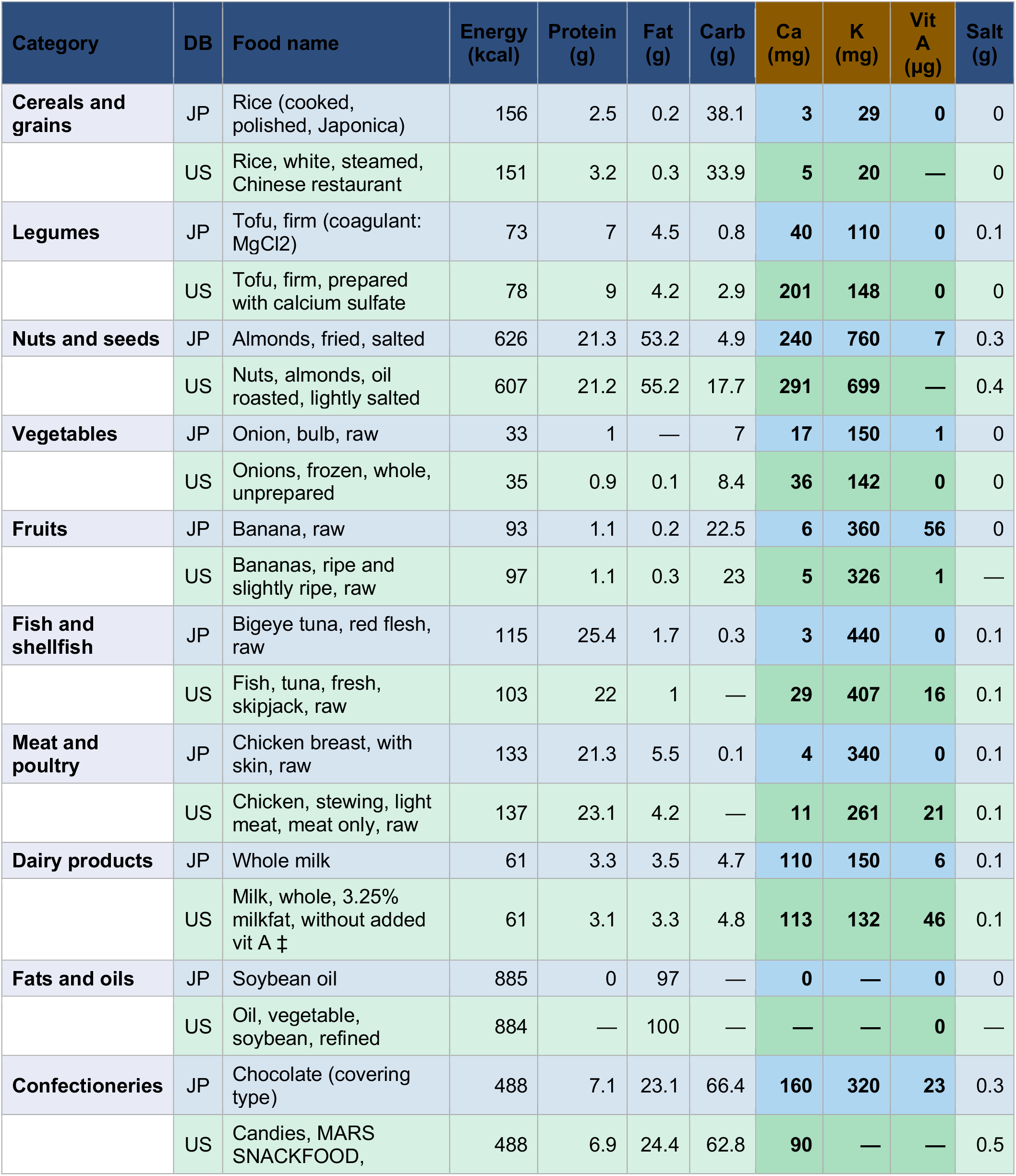
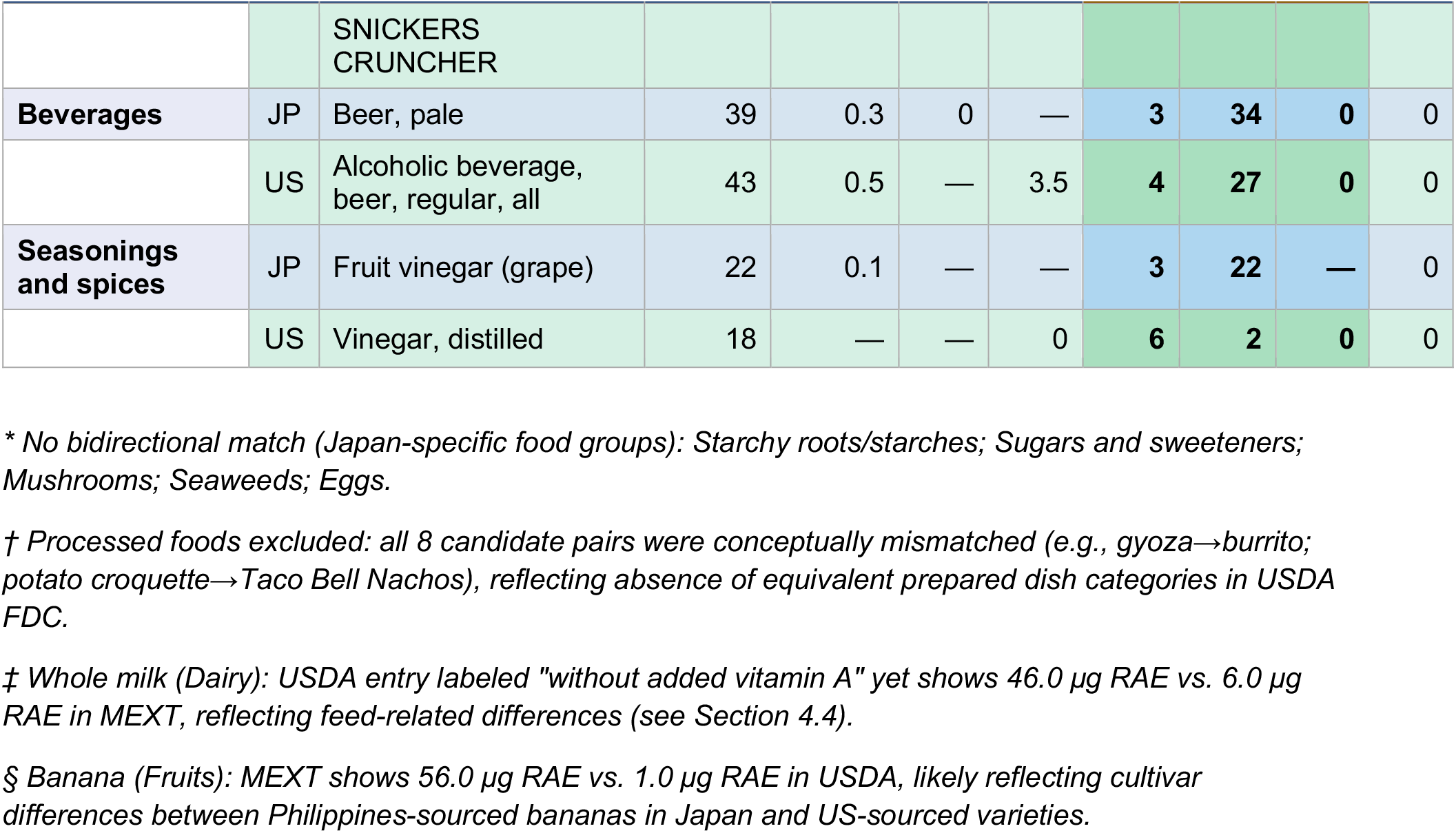
Representative bidirectional matched food pairs (one per MEXT category) with selected nutrient values per 100 g edible portion. JP = MEXT 2020 (8th ed.); US = USDA FoodData Central. Shaded columns (Ca, K, Vit A) correspond to nutrients showing systematic cross-national differences (Figure 3). The complete correspondence table (n=435) is available at https://doi.org/10.5281/zenodo.20103327.

### 3.3 Systematic differences in calcium, potassium, and vitamin A

Table 1 and Figure 3 present nutrient differences (FDC − MEXT) across food categories. At the aggregate level, FDC items showed higher energy (+2.2 kcal), carbohydrate (+2.6 g), and calcium (+6.0 mg), while MEXT items showed higher potassium (−3.7 mg) and vitamin A as RAE (−3.7 μg) per 100 g.

**Figure 3.**
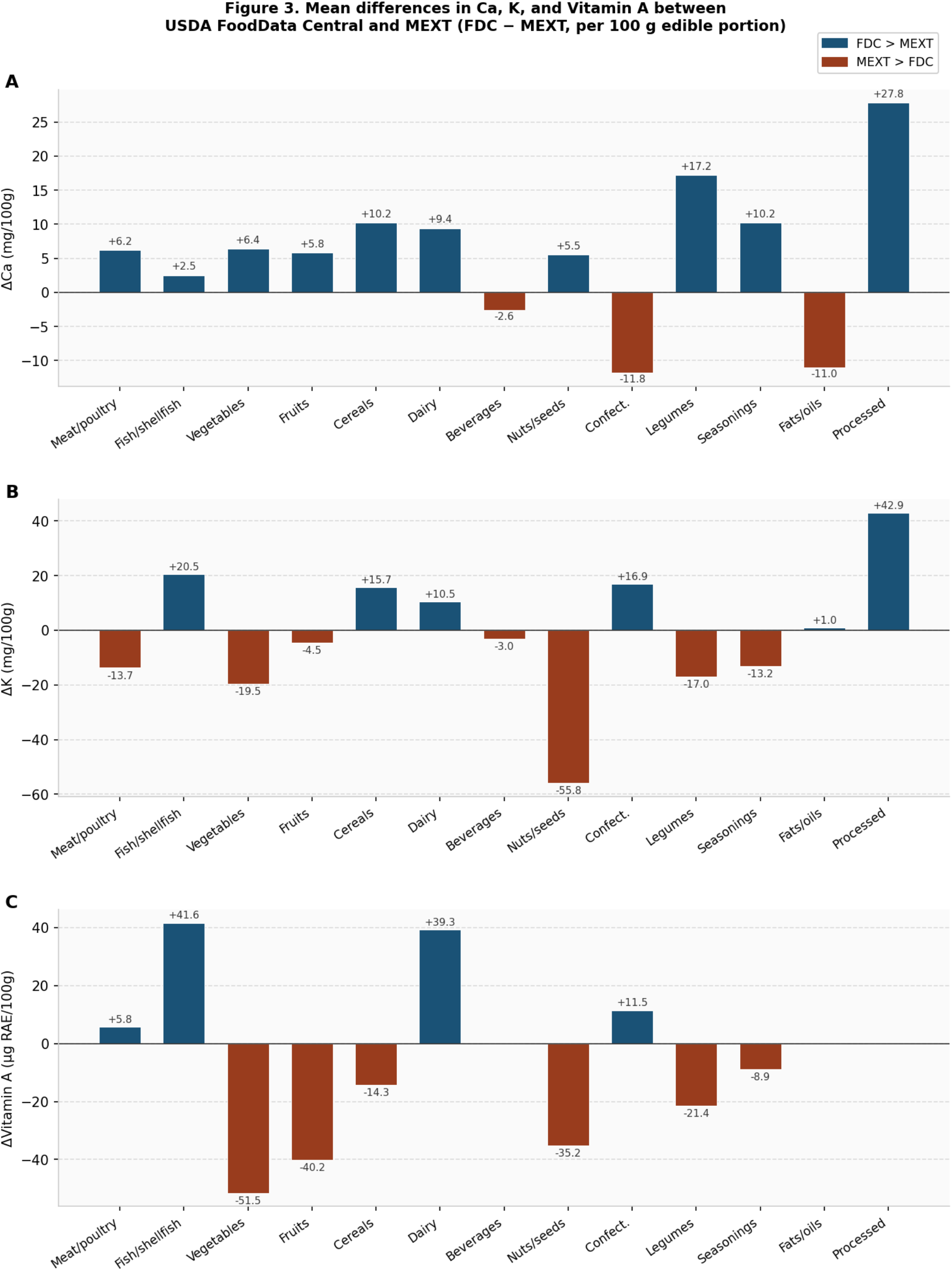
Mean differences in calcium (A), potassium (B), and vitamin A as RAE (C) between USDA FoodData Central and MEXT across 13 food categories (FDC − MEXT, per 100 g edible portion, n=435). Blue bars: FDC > MEXT; brown bars: MEXT > FDC. Outlier pairs (n=21) excluded.

The most striking finding was the directional consistency of differences in Ca, K, and vitamin A. Calcium was higher in FDC in 12 of 13 categories (Figure 3A), potassium was higher in MEXT in 9 of 13 categories (Figure 3B), and vitamin A was higher in MEXT in 8 of 13 categories (Figure 3C). This cross-category directional consistency strongly suggests that these differences reflect structural features of the food supply in each country rather than random matching error.

A particularly illustrative case is whole milk: the USDA entry explicitly states “without added vitamin A,” yet its vitamin A content (46.0 μg RAE/100g) is approximately 8-fold higher than the corresponding MEXT value (6.0 μg RAE/100g). This difference cannot be attributed to fortification and must reflect intrinsic differences in milk composition arising from differences in feeding practices. A notable exception in the fruit category was banana, where MEXT showed substantially higher vitamin A (56.0 μg RAE) compared to USDA FDC (1.0 μg RAE), likely reflecting differences in cultivar composition between Philippines-sourced bananas common in Japan and US-sourced varieties.

## 4. Discussion

### 4.1 A novel finding: systematic cross-national differences in Ca, K, and vitamin A

The principal novel finding of this study is the systematic, category-consistent directional difference in calcium, potassium, and vitamin A between matched Japanese and American food items. To our knowledge, this is the first time such a pattern has been documented at the food-item level across a comprehensive set of basic foodstuffs common to both countries.

The directional consistency of the calcium difference — positive (FDC > MEXT) in 12 of 13 food categories — is particularly compelling. If this pattern were due to matching errors, we would expect approximately equal numbers of positive and negative errors across categories. Instead, the near-universal elevation of calcium in FDC items relative to their MEXT counterparts suggests a systematic property of the American food supply.

### 4.2 Mechanisms underlying calcium differences

Several mechanisms may explain the systematically higher calcium in FDC items. First, calcium fortification of staple foods is substantially more prevalent in the United States than in Japan [8]. Second, drinking water hardness is considerably higher in many parts of the United States, contributing to dietary calcium through food preparation [9]. Third, US regulatory standards permit and encourage calcium fortification across a broader range of food categories than is customary in Japan [10].

The calcium elevation is observed even in categories such as meat and poultry (+6.2 mg) and fruits (+5.8 mg), where direct fortification is uncommon, suggesting that factors beyond targeted fortification — possibly feed composition for livestock, soil calcium content, and water used in food processing — may contribute to a baseline elevation across the American food supply.

### 4.3 Mechanisms underlying potassium differences

The systematically lower potassium in FDC items — most pronounced in nuts and seeds (−55.8 mg) and vegetables (−19.5 mg) — is particularly intriguing given the well-established importance of dietary potassium in cardiovascular protection [11].

Potassium is a water-soluble mineral that leaches readily during cooking. Japanese culinary traditions typically involve brief blanching followed by seasoning, preserving more cellular potassium than American cooking methods that may employ longer boiling times [12]. Additionally, Japan’s predominantly volcanic soils are characteristically rich in potassium, potentially resulting in higher baseline concentrations in domestically grown produce [13].

The practical implication is direct: researchers using FDC data to estimate potassium intake in Japanese populations will introduce systematic bias. Given the robust evidence linking dietary potassium to blood pressure and cardiovascular outcomes [11], this bias may affect the interpretation of cross-national epidemiological comparisons.

### 4.4 Mechanisms underlying vitamin A differences

Vitamin A differences showed a complex pattern: MEXT vegetables showed markedly higher vitamin A (−51.5 μg), while FDC fish and shellfish (+41.6 μg) and dairy products (+39.3 μg) showed higher values. The higher vitamin A in MEXT vegetables likely reflects the prominence of carotenoid-rich green-yellow vegetables in the Japanese diet: komatsuna, shungiku, and mitsuba are exceptionally rich in β-carotene [14].

A particularly instructive example is whole milk. The USDA entry is explicitly labeled “without added vitamin A,” yet contains 46.0 μg RAE/100g — approximately 8-fold higher than the MEXT value of 6.0 μg RAE/100g. This cannot be explained by fortification and instead reflects the higher β-carotene content of pasture-fed dairy cattle more common in the United States compared to the compound feed-based production systems more prevalent in Japan [15].

The banana exception (MEXT: 56.0 μg RAE vs. FDC: 1.0 μg RAE) illustrates cultivar-level variation: bananas consumed in Japan are predominantly imported from the Philippines, where certain Cavendish cultivars may have higher β-carotene content than varieties reflected in USDA values.

### 4.5 Implications for international nutritional epidemiology

The findings have direct methodological implications for cross-national dietary research. International comparisons of nutrient intake that rely on a single national food composition database will systematically misestimate intake in the non-reference country. For calcium, MEXT-based estimates will underestimate relative to FDC; for potassium and vegetable-sourced vitamin A, FDC-based estimates will underestimate relative to MEXT.

The complete correspondence table (Version 0.1, n=435 pairs) is openly available at https://github.com/shnkgw-rincom/jbfd-correspondence-table (DOI: 10.5281/zenodo.20103327). Users are invited to report errors via GitHub Issues; verified corrections will be incorporated in future versions. This study also demonstrates that LLM-based food matching is scalable to other national database pairs without human-curated training data.

### 4.6 Limitations

Several limitations should be noted. First, the LLM matching pipeline is not infallible: 21 outlier pairs were excluded, and residual conceptual mismatches may remain. Notably, all 8 candidate pairs in the processed foods category were conceptually mismatched (e.g., gyoza matched to burrito; potato croquette matched to Taco Bell Nachos), reflecting the fundamental absence of equivalent prepared dish categories in USDA FDC; this category was therefore excluded from Table 2. Second, FNDDS Survey data exclusion means composite dishes and restaurant items are not represented. Third, nutrient differences were analyzed per 100 g without accounting for typical serving sizes. Fourth, both databases represent national averages that may not capture regional or seasonal variation. Fifth, proposed mechanisms require direct experimental validation.

Subsequent papers will address: Japan-specific and US-specific foods (Paper 2); one-directional match interpretation and many-to-one mapping (Paper 3); methodological review and extension to other national databases (Paper 4); and the Japan Branded Food Database (Paper 5).

## 5. Conclusions

We have established the first bidirectional correspondence table between the Japanese Standard Tables of Food Composition 2020 (8th edition) and the USDA FoodData Central, comprising 435 confirmed food pairs validated by bidirectional LLM-based matching. The systematic, cross-category directional differences in calcium (FDC higher, 12/13 categories), potassium (MEXT higher, 9/13 categories), and vitamin A (MEXT higher for vegetables, FDC higher for dairy/fish) reflect genuine structural differences in the food supply of Japan and the United States. These findings have direct implications for cross-national dietary comparison studies. The complete correspondence table is openly available (DOI: 10.5281/zenodo.20103327) and LLM-based matching offers a scalable approach to global food composition database harmonization.

## Data Availability

All data produced in this study are openly available at https://github.com/shnkgw-rincom/jbfd-correspondence-table (DOI: 10.5281/zenodo.20103327) under CC0 1.0 Universal license.

## Acknowledgements

The authors thank Dr. Mieko Kimura, Director of the Takeda Life Science Research Center, for her long-standing guidance on the scientific insights underlying this research. The authors also thank the Ministry of Education, Culture, Sports, Science and Technology, Japan for providing MEXT 2020 (8th edition) in open-access format, and the USDA Agricultural Research Service for providing FoodData Central under CC0 1.0. LLM-based matching was performed using the Anthropic Claude API.

## Data Availability

All datasets generated in this study are openly available at https://github.com/shnkgw-rincom/jbfd-correspondence-table (DOI: 10.5281/zenodo.20103327) under CC0 1.0 Universal license. The repository contains five subset files: (1) bidirectional consensus pairs (correspondence_v0.1.csv, n=435); (2) JP→FDC matched pairs (jp_to_us_matched_v0.1.csv, n=1,927); (3) FDC→JP matched pairs (us_to_jp_matched_v0.1.csv, n=5,445); (4) Japan-specific foods with no FDC equivalent (jp_only_v0.1.csv, n=549); and (5) US-specific foods with no MEXT equivalent (us_only_v0.1.csv, n=2,698). Matching pipeline scripts are also publicly available in the same repository.

